# You aren’t what you eat, you become what you eat

**DOI:** 10.1101/2021.08.23.21262191

**Authors:** Christopher R. Stephens, Jonathan F. Easton, Heriberto Román Sicilia

## Abstract

Obesity (and the consequent obesity epidemic) is a complex, adaptive process, taking place over a time span of many years. Energy intake is recognized as a potentially important driver of obesity, especially in the context of an identifiable energy imbalance which, it is surmised, must lead to weight gain. Similarly, energy expenditure must play an important role. However, both show an enormous degree of individual variation. Therefore, measuring them is an exceedingly difficult task, especially in the context of large populations and long time periods. It has been argued that population-level observed weight gain can be traced back to very small daily energy imbalances while, at the same time, positing that a much larger maintenance energy gap is responsible for maintaining the energy requirements of the increased weight population. In this paper we examine the relation between BMI and energy intake as functions of age. The convexity of the BMI curves as a function of age and gender demonstrate the enhanced obesity risk apparent in young adults and women, and imply that no settling points exist at the population level. Consistent with other studies, overall weight increases are consistent with a very small daily energy imbalance, about 7 cal. Consumption as a function of age shows a small, steady, linear decrease of about 8 cal per year, and can be associated with a maximal energy excess/deficit of about 250cal for the youngest and oldest age groups. By examining weight differences between age groups as a function of age, we argue that this excess/deficit is an important motor for the observed weight differences, and argue that the apparent energy imbalance of 250 cal, due to excess consumption, leads to an effective imbalance of only 7 cal due to the existence of various physiological and behavioral mechanisms that enhance weight homeostasis and effectively reduce the energy excess from 250 cal to 7 cal. We discuss several possibilities for such mechanisms.

## 1. Introduction

It is gradually being recognised that standard reductionist approaches to the large worldwide increase in obesity levels are not working; neither from a practical perspective [1, 2], nor from a theoretical standpoint [3–6]. Complexity science can offer a different framework in which the problem of obesity may be studied [7–10]. The fact that obesity is a complex problem is manifest in its enormous multi-factoriality, where the individual risk factors cover a large spectrum of scales, ranging from the microscopic (such as genetics, and epigenetics [11, 12]), to the macroscopic (such as government policy and food industry marketing [13–14]).

Science is not accustomed to such systems, especially from a data point of view. Besides the fact that it is very multi-factorial, equally important is the fact that obesity is highly dynamic and highly influenced by *adaptive* changes, both in the individual and the environment. Unfortunately, our capacity to mathematically model Complex Adaptive Systems (CAS) is poor. In the absence of a methodological framework within which to do so, at the very least we should adopt a data perspective that respects the complex, adaptive nature of obesity and that allows us to develop a deeper phenomenological understanding of it from which quantitative models may be built. Put simply, the phenomenology of CAS is much richer than that of simpler systems. Even the case of, seemingly, one relationship, between Body Mass Index (BMI) and calorie consumption -the focus of this study- is enormously complex, and driven by a complex hierarchy of causal relations, many of which have caused much controversy, such as the “is a calorie a calorie?” debate [15–19].

The multifactorial, dynamic nature of the obesity epidemic as a CAS poses particular challenges from a data point of view. First of all, its multifactorial nature requires large populations with a large number of risk factors being recorded, so as to be able to represent many potential factors at a reasonable level of statistical significance. Secondly, it requires longitudinal studies over long time periods, in order to be able to observe adaptation taking place at distinct time intervals. Studies that combine a great deal of data depth, large populations and substantial time periods are obviously prohibitively difficult and expensive.

In the case of the relation between BMI and consumption there are myriad studies, of different types. Many are associated with controlled feeding studies [20–24], where the consumption pattern of a small population (∼ 10-100) is controlled over a relatively short time period (∼ 1–6 months) and related to BMI changes at the individual level. Such studies have been used, in conjunction with very detailed mathematical modelling of human metabolism [25–27], as the basis for extrapolations to much longer time periods. However, there is little to no evidence that physiological and behavioural adaptations on the scale of a month or so are analogous to what happens over a lifetime. In contrast, both transverse [28] and longitudinal [29] larger scale epidemiological studies have been used to study BMI and consumption [30–34]. The problem there is that inferences about the link between consumption and weight gain are much more indirect and, therefore, the distinction between correlation and causation is even more difficult. In the absence of large scale, highly multifactorial, longitudinal studies however, all current approaches require some degree of extrapolation.

A fundamental notion in considering the relation between BMI and consumption, is that of energy imbalance [35–44], the idea, with its root in thermodynamics, that weight gain has to be due to energy intake being greater than energy expenditure. Undeniable as this may be, there are a host of associated unknowns, the most fundamental being why energy consumption is greater than energy expenditure in the first place, either at the individual or the population level. It is not fully known how energy is harvested from food, both as a function of diet composition – protein versus fats versus carbohydrates – or of body state. For instance, it is now known that energy harvest depends on the composition of the microbiota [45–48]. In other words, the assumption that all the calorific content of ingested food is available for expenditure is not necessarily true and could also be dynamic and under some form of adaptive control. In addition, it is not known how much energy is expended, at least in realistic conditions and over longer time scales. Although, the doubly labelled water technique [49, 50] gives a good representation of the metabolic expenditure of energy it does not necessarily give a good account of other types of energy expenditure, such as changes in the amount of energy radiated to the environment through vasoconstriction and vaso-dilation.

Finally, it is not known for a given energy intake how to map this to energy expenditure in a way that includes all the above mentioned processes. The most advanced framework for doing so is that developed by Hall and collaborators [25–27], a tour de force that derives a phenomenological map between consumption, as broken down into proteins/lipids/carbohydrates, and changes in energy storage, given a certain body state in terms of the division into fat free and lean mass. The model is phenomenological, as some of its parameters are fitted to the results of the Minnesota starvation/refeeding experiment [20]. Our chief concern with such models is whether or not they reflect any mechanisms for long-term adaptive changes in human physiology, as opposed to just short-term changes, and to what extent they miss other mechanisms for energy expenditure and adaptation in energy harvesting, some possibilities of which are mentioned above.

In this paper, we will study the relation between BMI and consumption using data from the ENSANUT 2006 study [51, 52] cognizant of the fact that we will, like other epidemiologically based studies, be making strong assumptions and related inferences. We will consider consumption levels and BMI as a function of age and then, in distinction to other such studies, we will infer a relation between BMI differences between adjacent age groups and an associated consumption level. Consequent to this analysis we hypothesise that an age dependent excess/deficit of calories relative to a particular settling point (energy imbalance) is a prime motor for the convex nature of the relation between BMI and age.

The maximum inferred energy imbalance is about 250 cal, suggestively similar to the average increase in consumption of 252 cal over the period 1971-2000 as deduced from the NHANES data, and as reported in [36]. It is also similar to the proposed energy maintenance gap inferred in [35]. In purely energetic terms, the observed population level weight difference, interpreted as weight gain, is consistent with a very small daily energy imbalance of about 7 cal, as inferred from other studies [43]. We resolve the contradiction between an effective observed imbalance of 250 cal and an energetically self-consistent imbalance of 7 cal by hypothesising that there exist multiple physiological and behavioural mechanisms that maintain a much higher degree of homeostasis than would be expected given an energy intake excess of 250 cal. We discuss several of these possible mechanisms as fruitful ground for further research.

## 2. Materials and Methods

The data used is for 20,360 adults aged 20 or above who participated in the Mexican National Health and Nutrition Survey in 2006 [51, 52]. This survey used a stratified, multistage probability cluster sampling design to select participants of civilian, non-institutionalized populations. It contained a detailed nutritional questionnaire. The data was categorised into three categories – all adults, male and female. Exclusions were based on incomplete data for the purpose of this study, presence of diabetes, pregnancy, total equivalent reported caloric intake < 500 cal or > 5,000 cal or age > 80.

The ENSANUT 2006 survey was a food frequency questionnaire covering 101 different food items. As is known, any methodology for self-reporting of food intake is subject to reporting bias, with each methodology having advantages and disadvantages [53, 54]. However, despite these defects, food frequency questionnaires are still currently a primary way to assess usual dietary intake, and thus, the dietary habits of a population [55–58]

For each food item considered in the survey there are corresponding questions for the number of days per week and number of times per day the item was consumed within the seven days before the interview date. Also included was the portion size (weight) and number of portions. The food table included in the questionnaire, as well as the portion size (very small, small, medium, large, and very large) and mean weight of portion, for each population group, were estimated by a group of researchers from the Instituto Nacional de Salud Pública (INSP) from an analysis of the most-consumed-foods data obtained from the National Nutrition Survey 1999. We calculated per-week and per-day quantities of each food item consumed using the number of days per-week, times per day, portion size (weight) and number of portions responses. The number of times per day variable was not included in our analysis, as in a test of the average for 20 randomly selected foods, across the four possible responses (1, 2-3, 4-5 or 6 times per day) 91.63% of participants responded eating the food just once a day. A similar analysis was also carried out for the size of portion question. The percentage of extra-small, small, medium, large and extra-large portions were taken from every food type with a response for this question. It was found that 93.2% of all 142,176 responses for portion size were medium. Also, the size of portion question was not included in the questionnaire for all food types. In conclusion, food consumption was determined from just the frequency per week and number of portions variables. Foods were categorized into 8 groups: Vegetables; Dairy products; Cereals and grains; Fresh fruit; Fish, Meat, Eggs and dry legumes; Sugars and fats; and Fast food. The food items contained in each food category can be seen in the Supplementary Material. Total daily consumption in a given food group was calculated by summing (number of times per week * number of portions)/7 over all the foods in a given food category. Total food consumption was then calculated by summing the portions consumed across all food groups.

The average total caloric intake for each person in the study was subsequently determined by computing the average caloric content of a portion in each food group and summing over the portions consumed in each group. The caloric content of a portion for each food group, except Fast food, was calculated by using averages provided in the tables of “Sistema mexicano de alimentos equivalentes” [59]. Due to the different taxonomy of food types, and the different portion sizes used in these tables versus the ENSANUT study, it was necessary to use a multiplier as a scale factor for each food group in order to give a good approximation to the average calories in relation to one portion of each food group.

The multiplier was calculated by comparing total caloric intake to the recommended level of calorie counters that are based on the base metabolic rate suggested from the Mifflin-St. Jeor equation [60]. Through testing, it was determined that a multiplicative scale factor of three gave an acceptable range of daily total calorie intake for each food group and for the total calories consumed. The average caloric content of the Fast food category was calculated by taking the average calories obtained from the United States Department of Agriculture (USDA) Agricultural Research Service [61], for each of the foods contained in the group as seen in Table 6 (Fast Food) of the supplementary material.

### Statistical Analysis

We performed linear regressions using SPSS of age, t, against BMI, and age against total calories consumed daily, C, for the three different groupings: all adults, male and female with sample sizes: 15,738, 5,662 and 10,076 respectively.

To consider changes in BMI as a function of age we considered the difference in average BMI between ages t and t+1, Δ(t) = (<BMI(t+1)>-<BMI(t)>), where <BMI(t)> is the average BMI for the participants of age t in the group of interest. As the data is not longitudinal, the populations for each age are not the same and, hence, to smooth the fluctuations due to this heterogeneity we considered a centralized moving average of Δ(t), Δ_MA_(t), defined as Δ_MA_(t) = (Δ(t-1)+ Δ(t)+ Δ(t+1))/3. We considered various other smoothing algorithms, such as regressing (Δ(t)+ Δ(t+1))/2 against (C(t)+C(t+1))/2 and other definitions of the moving average, with no corresponding significant change in our results.

We further considered regressions of Δ(t) and Δ_MA_(t) versus t, and Δ(t) and Δ_MA_(t) versus <C(t)>, where <C(t)> is the average reported calorie consumption per day for participants of age t in the group of interest. In this case the sample size is 58, corresponding to (age_max – age_min -2). In all cases, regressions were tested using only a linear term and with both a linear and quadratic term. The regression with the most significant f statistic was chosen and is shown in the results section.

For each regression we show the unstandardized regression coefficients, the corresponding standard errors and t statistics and the lower and upper bounds associated with the 95% confidence interval for the coefficients. We also show the f statistic, the R^2^ value and the significance of each regression.

## 3. Results

In Table 1 we see the number and percentage of participants by age interval and category in the sample. The larger number of female respondents is due to the fact that women were more likely to be at home when the interviewer called.

**Table 1:** Number and percentage of the different categories by age group.

| Gender | Male |  |  | Female |  |  | All Adults |  |
| --- | --- | --- | --- | --- | --- | --- | --- | --- |
|  | # | % Males | % Age | # | % Females | % Age | # | % Adults |
| Age |  |  |  |  |  |  |  |  |
| 20-29 | 1170 | 20.66% | 33.15% | 2359 | 23.41% | 66.85% | 3529 | 22.42% |
| 30-39 | 1511 | 26.69% | 31.70% | 3256 | 32.31% | 68.30% | 4767 | 30.29% |
| 40-49 | 1250 | 22.08% | 37.63% | 2072 | 20.56% | 62.37% | 3322 | 21.11% |
| 50-59 | 755 | 13.33% | 41.26% | 1075 | 10.67% | 58.74% | 1830 | 11.63% |
| 60-69 | 545 | 9.63% | 43.15% | 718 | 7.13% | 56.85% | 1263 | 8.03% |
| 70-80 | 431 | 7.61% | 41.97% | 596 | 5.92% | 58.03% | 1027 | 6.53% |
| Total | 5662 |  |  | 10076 |  |  | 15738 |  |

In Figure 1 we see a graph of BMI versus age for the 15,738 included participants. Also included is the data corresponding to average BMI, <BMI(t)>, by age, and a quadratic polynomial fit to the binned data. A linear fit was also considered but was less statistically significant.

**Figure 1:**
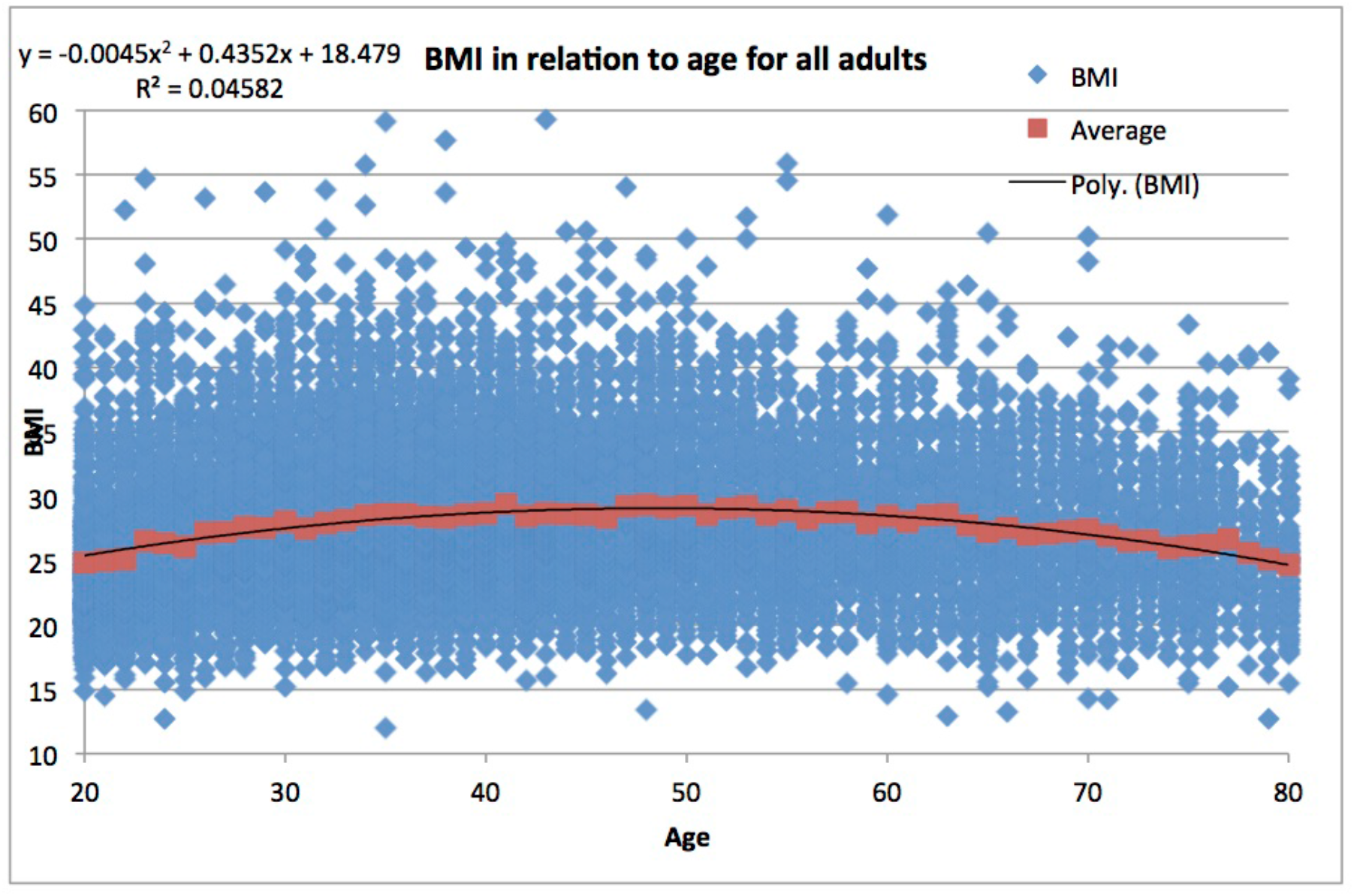
BMI against age for all adults.

The summary statistics for this regression can be seen in Table 2, where we see the relationship between age and BMI for the full sample, and for the different groupings, using a regression with linear and quadratic age terms. As can be seen, the fit to a quadratic curve is very impressive, with f-statistic values in the range 290-370 and absolute t values for the regression coefficients between 14 and 27. The relatively low value of the R^2^ coefficient is associated with the fact that, as can be seen in Figure 1, although the quadratic tendency is very statistically significant, there is also a great deal of underlying statistical variation.

**Table 2:** Regressions of BMI against age for all adults, male and female.

|  | Variable(s) | Unstd. B | Std. Error | t | f | R^2 | Sig | Lower | Upper |
| --- | --- | --- | --- | --- | --- | --- | --- | --- | --- |
| BMI |  |  |  |  | 372.668 | 0.045 | 0 |  |  |
| ALL | Constant | 18.533 | 0.347 | 53.445 |  |  | 0 | 17.853 | 19.212 |
|  | Age | 0.433 | 0.016 | 27.278 |  |  | 0 | 0.402 | 0.464 |
|  | Age^2 | -0.004 | 0 | -26.678 |  |  | 0 | -0.005 | -0.004 |
|  | Variable(s) | Unstd. B | Std. Error | t | f | R^2 | Sig | Lower | Upper |
| BMI |  |  |  |  | 103.539 | 0.035 | 0 |  |  |
| Men | Constant | 20.06 | 0.493 | 40.666 |  |  | 0 | 19.093 | 21.027 |
|  | Age | 0.321 | 0.022 | 14.347 |  |  | 0 | 0.277 | 0.364 |
|  | Age^2 | -0.003 | 0 | -14.326 |  |  | 0 | -0.004 | -0.003 |
|  | Variable(s) | Unstd. B | Std. Error | t | f | R^2 | Sig | Lower | Upper |
| BMI |  |  |  |  | 290.452 | 0.055 | 0 |  |  |
| Women | Constant | 17.399 | 0.46 | 37.821 |  |  | 0 | 16.497 | 18.301 |
|  | Age | 0.504 | 0.021 | 23.794 |  |  | 0 | 0.463 | 0.546 |
|  | Age^2 | -0.005 | 0 | -22.767 |  |  | 0 | -0.006 | -0.005 |

In Figure 2 we see a graph of average daily caloric consumption versus age for the 15,738 included participants. Also included is the data corresponding to average calorie consumption, <C(t)>, by age and a linear polynomial fit to the binned data. A quadratic fit was also considered but did not lead to a more statistically significant f value.

**Figure 2:**
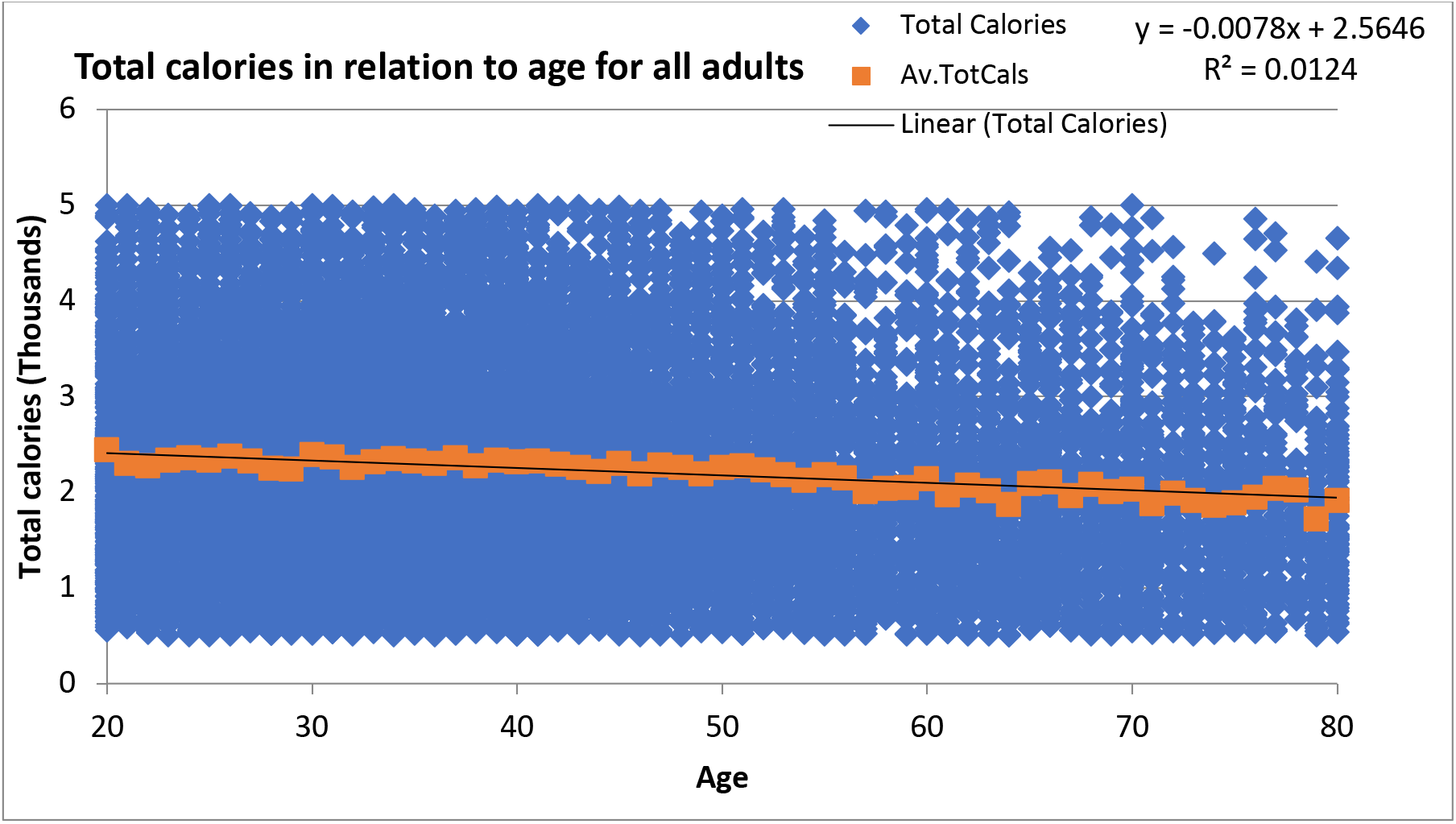
Daily calorie consumption against age for all adults.

In Table 3 we see the summary statistics for the relationship between age and total calorie consumption for the full sample and for the different groupings, using a regression with only a linear term. Once again, the strong statistical significance of the underlying relationship is apparent, with f values in the range 69 to 197, and absolute t values for the regression coefficients in the range 8-14. As with BMI, the low R^2^ value is an indication of a high degree of statistical variability.

**Table 3:** Regressions of total reported daily consumption against age for all adults, male and female.

|  | Variable(s) | Unstd. B | Std. Error | t | f | R<sup>2</sup> | Sig | Lower | Upper |
| --- | --- | --- | --- | --- | --- | --- | --- | --- | --- |
| Total Cals |  |  |  |  | 197.52 | 0.012 | 0 |  |  |
| ALL | Constant | 2.565 | 0.024 | 105.479 |  |  | 0 | 2.517 | 2.612 |
|  | Age | -0.008 | 0.001 | -14.054 |  |  | 0 | -0.009 | -0.007 |
|  | Variable(s) | Unstd. B | Std. Error | t | f | R<sup>2</sup> | Sig | Lower | Upper |
| Total Cals |  |  |  |  | 69.552 | 0.012 | 0 |  |  |
| Men | Constant | 2.638 | 0.042 | 62.809 |  |  | 0 | 2.556 | 2.721 |
|  | Age | -0.008 | 0.001 | -8.34 |  |  | 0 | -0.009 | -0.006 |
|  | Variable(s) | Unstd. B | Std. Error | t | f | R<sup>2</sup> | Sig | Lower | Upper |
| Total Cals |  |  |  |  | 144.087 | 0.014 | 0 |  |  |
| Women | Constant | 2.544 | 0.03 | 85.301 |  |  | 0 | 2.485 | 2.602 |
|  | Age | -0.008 | 0.001 | -12.004 |  |  | 0 | -0.01 | -0.007 |

Turning now to the relation between BMI and daily calorie consumption, in Figure 3 we see a graph of this relation for the 15,738 included participants. Average daily caloric intake was also binned into intervals of 100 cals over the range 500-5000 cal. The graph shows the relation between average BMI, <BMI(t)>, as a function of the binned average calorie consumption and a quadratic polynomial fit to the binned data. A linear fit was also considered but had a statistically less significant f value. Note the much smaller values of R^2^ for this regression compared to those of Figures 1 and 2.

**Figure 3:**
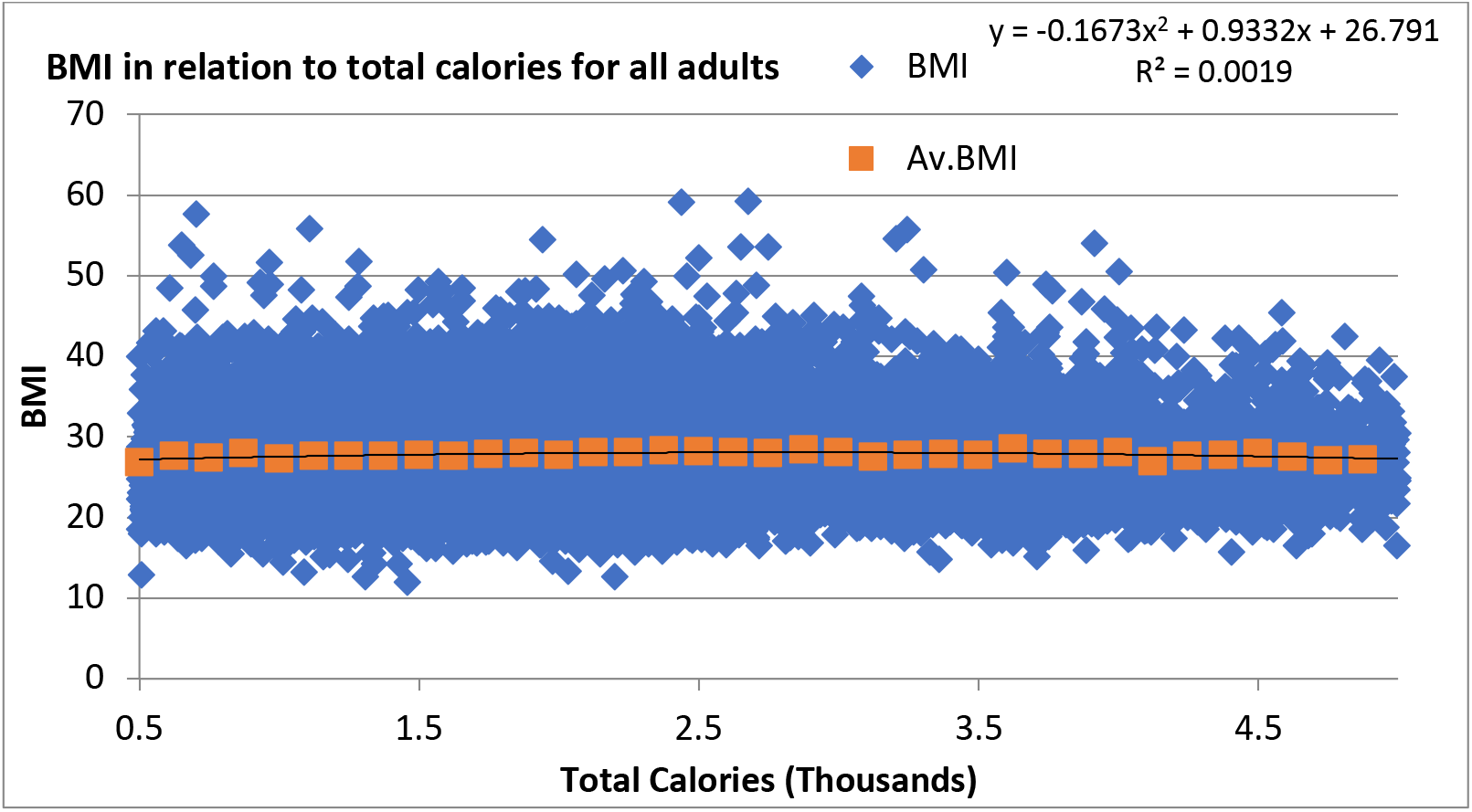
BMI against daily calorie consumption for all adults.

In Table 4 we see the summary statistics for the regression of BMI on total calories consumed daily for the full sample and for the different groupings. In this case, in contrast to the relationships between BMI and age, and consumption and age, and as observed in Figures 1-3, we see only a weak relation between BMI and consumption, with f statistic values that are approximately 15, 2 and 20 times less than the corresponding regressions for calorie consumption against age for all, men and women respectively. Note that, although a linear fit was found to be more appropriate for men and a quadratic fit for women, the impact of the quadratic term, as can be seen in Figure 3, is minimal.

**Table 4:**
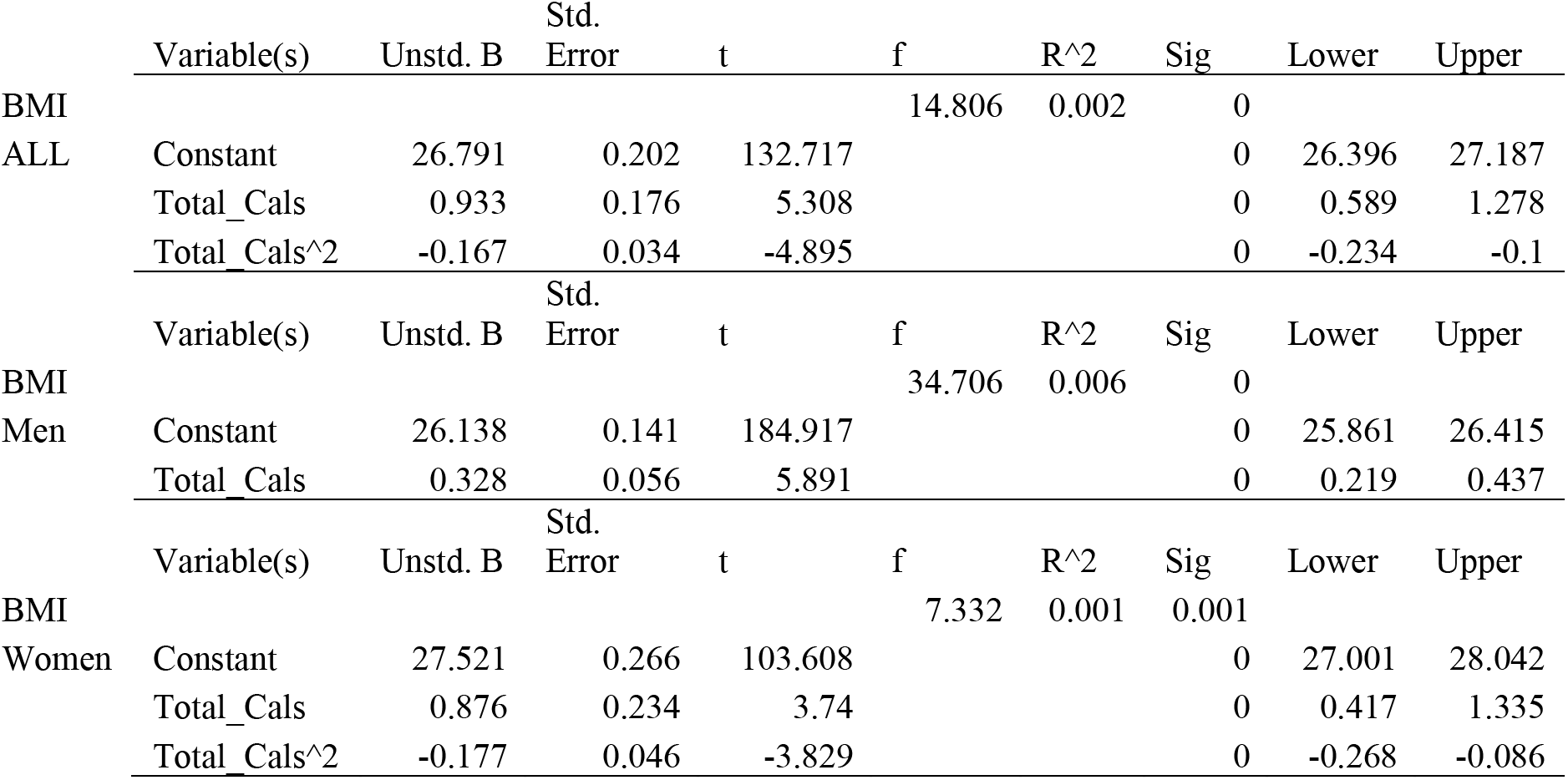
Regressions of BMI against reported daily consumption for all adults, male and female.

For BMI, calorie consumption and age it is possible to assign a value to an individual study participant. As the data is transversal however, it is not possible to assign a specific weight increase or decrease to an individual. For BMI changes therefore, we consider comparing average BMI in one age group versus another. This changes the sample sizes from 15,738, 5,662 and 10,076 for all, men and women respectively, to 58, corresponding to the independent age increments between ages 20 and 80 using a central moving average of BMI change. As might be expected, this potentially reduces the f values due to the large decrease in sample size, while at the same time allowing for substantial increases in R^2^ values as the variance is substantially reduced by the smoothing process.

In Table 5 we see the summary statistics for the relationship between change in average BMI, Δ_MA_(t), and age, t. The regressions for all categories are statistically significant. As anticipated, the smoothing leads to a decrease in the values of f and a strong increase in R^2^. For all three categories it was found that a regression with only a linear term was better. The results show that average annual BMI changes decrease linearly as a function of age, decreasing by a factor of approximately 0.1kg/m^2^ each year.

**Table 5:** Regressions of centralized moving average of year-on-year BMI change, Δ_MA_(t), versus age for all adults, male and female.

|  | Variable(s) | Unstd. B | Std. Error | t | f | R<sup>2</sup> | Sig | Lower | Upper |
| --- | --- | --- | --- | --- | --- | --- | --- | --- | --- |
| BMI Change ALL |  |  |  |  | 45.89 | 0.45 | 0 |  |  |
|  | Constant | 0.513 | 0.08 | 6.428 |  |  | 0 | 0.353 | 0.674 |
|  | Age | -0.01 | 0.002 | -6.774 |  |  | 0 | -0.013 | -0.007 |
| BMI Change Men |  |  |  |  | 14.05 | 0.201 | 0 |  |  |
|  | Constant | 0.387 | 0.11 | 3.531 |  |  | 0.001 | 0.168 | 0.607 |
|  | Age | -0.008 | 0.005 | -3.748 |  |  | 0 | -0.012 | -0.004 |
| BMI Change Women |  |  |  |  | 41.91 | 0.428 | 0 |  |  |
|  | Constant | 0.595 | 0.097 | 6.163 |  |  | 0 | 0.402 | 0.789 |
|  | Age | -0.012 | 0.002 | -6.473 |  |  | 0 | -0.015 | -0.008 |

In Table 6 we see the relationship between change in average BMI, Δ_MA_(t), versus <C(t)>. Once again, regressions with only a linear term were found to yield more statistically significant results. The regressions are again statistically significant for all three categories. The results show that average annual BMI changes increase linearly as a function of calorie consumption, increasing by about 0.8 kg/m^2^ per increase of 1000 calories.

**Table 6:** Regressions of centralized moving average of year-on-year BMI change, Δ_MA_(t), against average reported daily calorie consumption, <C(t)>, for all adults, male and female.

|  | Variable(s) | Unstd. B | Std. Error | t | f | R<sup>2</sup> | Sig | Lower | Upper |
| --- | --- | --- | --- | --- | --- | --- | --- | --- | --- |
| Moving Av. |  |  |  |  | 29.236 | 0.343 | 0 |  |  |
| BMI Change | Constant | -1.954 | 0.362 | -5.392 |  |  | 0 | -2.68 | -1.228 |
| ALL | Total Cals | 0.904 | 0.167 | 5.407 |  |  | 0 | 0.569 | 1.239 |
|  | Variable(s) | Unstd. B | Std. Error | t | f | R<sup>2</sup> | Sig | Lower | Upper |
| Moving Av. |  |  |  |  | 13.397 | 0.193 | 0.001 |  |  |
| BMI Change | Constant | -1.625 | 0.444 | -3.656 |  |  | 0.001 | -2.515 | -0.734 |
| Men | Total Cals | 0.724 | 0.198 | 3.66 |  |  | 0.001 | 0.328 | 1.121 |
|  | Variable(s) | Unstd. B | Std. Error | t | f | R<sup>2</sup> | Sig | Lower | Upper |
| Moving Av. |  |  |  |  | 22.429 | 0.286 | 0 |  |  |
| BMI Change | Constant | -1.754 | 0.372 | -4.711 |  |  | 0 | -2.5 | -1.008 |
| Women | Total Cals | 0.833 | 0.176 | 4.736 |  |  | 0 | 0.481 | 1.185 |

## 4. Discussion

We have emphasized that the obesity epidemic is both complex and adaptive. Its complexity is manifest in its multi-factoriality, with risk factors ranging from the micro to the macro. In the present context, this multi-factoriality is, in turn, reflected in the high degree of variance seen in Figures 1-3. In other words, the diffuseness of the “cloud” is not associated with statistical (read unpredictable) noise but, rather, with the complex heterogeneity that exists between individuals. A consequence of this heterogeneity is the difficulty of predicting at the individual level the relation between consumption and weight. However, despite this heterogeneity there are very robust, statistically significant, global features in the data that characterise the relationship between BMI, self-reported energy intake and age.

The salient facts are the following: i) BMI, or weight, follow a parabolic trajectory as a function of age; ii) self-reported energy intake is a linearly decreasing function of age; iii) BMI is approximately constant as a function of energy intake when considered across all ages.

The convexity of the relationship between age and BMI implies that the highest degree of weight increase/decrease is among the youngest/oldest adults. This convex relationship is a robust feature of the relation between BMI and age and also appears in data associated with multiple other studies [31, 62], including longitudinal ones [63]. In this particular data set, it manifests itself in a remarkably simple way, as seen in Table 2, where we see that the quadratic relationship between BMI and age is extremely statistically significant.

The convex, non-linear nature of the relation between BMI and age in itself has several important implications. Firstly, how obesity risk changes as a function of age: young adulthood, 20-30, is a particularly crucial time, in which there is an accelerated weight increase [62, 63] and at which public health initiatives should be directed. The expected increase in BMI from age 20 to 25 is 1.27 kg/m^2^, corresponding to an increase of 5% relative to the age 20 BMI, while from age 45 to 50 it is 0.27 kg/m^2^, corresponding to an increase of only 0.9% over the age 45 BMI. More than 27% of total weight change is attributable to that initial 5-year period, while for the period 45-50 it is less than 6%. By gender, the total increase in BMI from age 20 to peak BMI is 3.37 kg/m^2^ for men and 4.62 kg/m^2^ for women. The increases from age 20 to 25 are 27.6% of total weight increase for men and 30.1% for women. Similarly, the maximum decrease in BMI is from age 75-80, and is the same as the corresponding increase from 20-25. The maximum BMI for the three groups occurs at ages 54.1, 53.5 and 50.4 for the all, men and women groups respectively, with corresponding maximal BMI of 30.25, 28.6 and 30.1.

The symmetry of the relation between BMI and age is quite remarkable. Perhaps even more so given that the data is transversal. Of course, there are several factors that should be accounted for when trying to interpret the age dependence of BMI. First of all, as ENSANUT is a transversal study, although the present social and economic environment for a 20-year-old and an 80-year-old is approximately the same, the environment at age 20 of the 80-year-old would have been substantially different. In other words, the group of 80- year-olds have a 60-year life history inherent in their BMI distribution. Secondly, there is a potential selection effect, in that, in a strictly longitudinal, cohort study, some participants would have died, and one would expect an enhanced mortality rate for the morbidly obese.

We have examined this hypothesis in the context of NHANES data [28]. However, the contribution to the decrease in average BMI associated with potentially enhanced mortality of the obese is substantially smaller than the observed overall effect here. This means that an explanation of the decrease in BMI beyond age 50 involves multiple factors. Indeed, in geriatric studies [64–66] a decrease in calorie consumption, particularly associated with reduced protein consumption, has been noted but not explained [67–70]. In addition, the point of maximal BMI occurs at about age 50, much younger than where one would expect enhanced mortality to play a significant role. In other words, the fact that the population average of BMI begins to decrease after age 50 must be explained by factors other than a survivor bias.

Turning now to the relation between age and total energy intake, the fit to a linear model, seen in Table 3, is extremely statistically significant though, as with the BMI-age relation, it manifests a high degree of variance. The reason for this is analogous to that of BMI itself, in that there is a high degree of multi-factoriality associated with consumption levels. From the fitted relation, we may deduce that maximum average caloric intake, for both genders, occurs at age 20, and the minimum at age 80. The maximum average calorie intake across all participants is 2,565 cal at age 20. For men it is 2,638 cal and for women 2,544 cal. All groups are consistent with an average yearly decrease in daily energy intake of approximately 8 cal. This, of course, is an exceedingly small number, corresponding to a 0.32% reduction in daily calorie intake per year, relative to the male daily average, and a 0.3% reduction relative to the age 20 daily maximum. However, it corresponds to an important decrease in overall energy intake of approximately 500 cal when aggregated between age 20 and age 80. For women, the average yearly reduction is 0.4% of average daily caloric intake and 0.31% relative to the age 20 daily maximum.

Considering the relation between BMI and calorie consumption, as seen in Table 4: Compared to the results of Tables 2 and 3 the statistical relationship between BMI and consumption is much weaker than that between age and BMI, or age and energy intake. For all participants and women, the f and R^2^ statistics are about an order of magnitude less, and the t statistics for the regression coefficients about 5 times smaller. For men, although there is a notable drop in the statistical significance of the BMI versus energy intake relation relative to the BMI versus age and consumption versus age relations, there still appears to be an underlying correlation between BMI and energy intake. However, the generally much weaker relationship between BMI and energy intake is consistent with the conclusion that high BMI individuals do not consume more than low BMI individuals when considered across all age groups. In other words, this evidence suggests that: “you are not what you eat”. Note that this conclusion does not imply that an obese individual of a given age consumes less than a normal weight individual.

### BMI Changes as a Function of Consumption

If we accept that it is energy imbalance that causes weight gain, a natural hypothesis for the relation between BMI and energy intake can be deduced from Tables 2 and 3, if we take the maximum of the parabola to be the energy balance point, where energy intake does not change BMI. In this case, relative to this energy balance point of the distribution at age t_b_, for age t < t_b_ we may define an “excess” energy intake, while, for t > t_b,_ we may define an energy “deficit”, with the corresponding maximum energy intake excess/deficit being given by C(t = 20) and C(t = 80) respectively. Thus, the quadratic nature of the BMI vs age distribution would imply that there exists a positive energy imbalance up until age, t_b_, corresponding to the maximum, where energy imbalance is inferred to be zero, as there is no change in BMI, followed by a negative energy imbalance. Moreover, the curve implies that, at least at the population level, there is no settling point per se [71] but, rather, a continuous change in BMI as a function of age.

This, in turn, leads us to consider differences in BMI, rather than BMI itself, across the population as a function of age. In Table 5 we see the results of a regression of BMI change against age. The relevant statistical diagnostics must now be interpreted in the context of a statistical sample of 58 data points as opposed to 15,738. The maximum BMI change is predicted to occur between age 20 and age 21, with corresponding values of 0.513 kg/m^2^ (about 1.23kg) for all participants, 0.387 kg/m^2^ (about 1kg) for men and 0.595 kg/m^2^ (about 1.36kg) for women if we consider the average height in each group. This is consistent with the interpretation that, in this population, year on year changes in BMI and weight for women are substantially higher than those for men. The age at which BMI changes are zero is 51.3 for the group of all participants, 49.6 for women and 48.4 for men. These are generally consistent with the ages - 54.1, 50.4 and 53.5 for all, women and men respectively - associated with the maximum of the BMI versus age models, t_b_. Beyond these ages, BMI changes are negative, corresponding to a decrease in average BMI as a function of age with the maximum yearly decrease at age 80.

Note that this interpretation also gives a natural explanation for why there is no significant increase in BMI as a function of energy intake in Figure 3. Referring to Figure 2, due to the symmetry associated with the BMI vs age distribution, a given average BMI occurs at two symmetrically distributed ages. For instance, average BMI = 25 occurs at age 20 and age 80. However, at those symmetrical points the energy intake has equal contributions from an excess and a deficit, such that the sum is approximately zero. In other words, the data is consistent with the observation that the population characterised by a given average BMI is composed of two sub-populations, one younger that has an excess energy intake, and an older one that has an energy deficit. Of course, in this conclusion we are not stating that BMI does not increase for an individual as a function of energy imbalance; as, for example, in overfeeding. Rather, at the population level we are observing that BMI is not a single-valued function of energy intake.

We hypothesize then that the observed increase and subsequent decrease in BMI seen in Figure 2 is generated by an energy imbalance associated with an excess and subsequent deficit of calories, as seen in Figure 3. This hypothesis is examined and quantified in Table 6, where we see that the change in BMI is linearly correlated with calorie consumption. From Table 6 we may deduce that, for the all participants group, 2,162 cals is the reported consumption level that corresponds to zero increase in BMI. For men, the corresponding level is 2,244 cals, and for women 2,106 cals. With the relationship between BMI change and reported calorie consumption in hand that is consistent with the data, we may deduce what excess consumption leads, on average, to a given BMI increase. For instance, for women, a yearly increase in BMI of one kg/m^2^, corresponding to a 2.6 kg weight gain per year for an average height woman, implies a daily dietary intake of 3,306 cals, compared to a 2,106 calorie no BMI increase base level, or a recommended intake of approximately 2,000 cals. This corresponds to a 57% increase in calorie consumption. A BMI change of 1 kg/m^2^ every two years, corresponding to passing from overweight to obese in 10 years, corresponds to a calorie consumption of 2,706 cals, an approximately 28% consumption increase over the zero BMI change rate. However, in Figure 1 and Table 1 we observe that average BMI change, as a function of age, is not large, corresponding to a change in average BMI from overweight to obese over a period of 30 years. The reported consumption level consistent with this BMI growth level for women is 2,306 cals per day, only 200 cals (9.5%) per day more than the zero BMI change level. For men, a BMI change of 1 kg/m^2^ per year, i.e., on average a 3kg per year weight change, corresponds to a consumption level of 3,626 cals compared to the 2,244 zero BMI change rate, i.e., a 61% increase in caloric intake relative to the zero BMI change level. A BMI change of 1 kg/m^2^ every two years corresponds to a consumption level of 2,935 cals, and a change from overweight to obese (change from BMI = 25 to BMI = 30) over a 30-year period, as seen in the data, to a daily intake of 2,475 cals, 10% larger (230 cal) than the zero BMI change intake level.

As well as determining what energy intake corresponds to a given BMI change, it is also useful to determine the effect of a given fixed increment in energy intake. For men, for every thousand cal increase in daily consumption, BMI increases by 0.72 kg/m^2^ per year, on average about 1.94kg, while, for women, the corresponding increase is 0.83 kg/m^2^, 15% greater than the corresponding male value, but also corresponding to a similar weight increase of about 1.89kg. The maximal energy excess of 268/243 cal per day for men/women at age 20 corresponds to a BMI increase of 0.19 kg/m^2^ and 0.20 kg/m^2^, corresponding to a weight gain per year for average size men/women of 0.53/0.46 kg respectively. Note that this energy “gap” has been derived at the population level and is not the same as the standard energy gap associated with the difference between daily energy intake and expenditure for an individual [35–37].

It is interesting, however, that the inferred motor for BMI differences here is about the same as the maintenance energy gap/energy flux gap [25, 43, 72] that has been derived from comparing the overall weight gain in the US population over a 30-year period, to the different energy requirements of the average population weights at the two extremes of this period, as derived from studies that used the doubly-labelled water technique to measure total energy expenditure versus weight. This analysis relies on the assumption that on a daily basis each individual is in approximate energy balance and that the end points of the analysis can be identified as “settling points”. As emphasized by Schutz et al [36], however, in reality this is not so much an energy “gap” related to the notion of an imbalance that can be interpreted as a driver of weight change but, rather, an obligatory consequence of the existence of a true energy gap/imbalance. In contrast, the excess consumption noted here we do interpret as a true energy imbalance and the driver of population level BMI changes.

It is important to emphasize that the observed and postulated relationships between energy intake and BMI do not offer a “complete” explanation but, rather, offer evidence that pure caloric consumption is, indeed, an important quantifiable motor for BMI increase. The large variance inherent in the relationship is evidence of the complex, multi-factorial nature of the obesity epidemic and indicates the importance of adopting a multi-factorial modelling approach in order to determine what part of this variance is predictable through the effect of other measurable variables and what part is irreducible.

Thus, we conclude that the observed distributions of BMI and energy intake are consistent with the hypotheses that: i) excess energy intake is an important driver of BMI increases; ii) that this energetic excess is a decreasing function of age, being a maximum/minimum at age 20/80 (for this population); and iii) that the relative excess for a given age is substantially greater for women than men. The overall conclusion can be phrased thus: it’s not that “you are what you eat”, but more that “you become what you eat”.

### Population Energy Balance as an Emergent Phenomenon

An important lesson to draw from our analysis is that associated with the difficulty of separating signal from noise. The data is consistent with the hypothesis that, at the population level, there is an average yearly decrease of about 8 calories in daily consumption. However, the standard deviation of the daily consumption for this population is about 1300 cal. Even the maximum daily calorific excess of 200-300 cal is small compared to this number. Additionally, the average increase in BMI from age 20 to age 50 is about 0.15 kg/m^2^ per year, corresponding to an average weight gain of about 400g per year. Compared to the standard deviation in BMI over the population of 5.17 kg/m^2^ this is also a very small amount. This shows the difficulties of determining the relationship between consumption and BMI change at the individual level under realistic conditions, i.e., not with intense overfeeding or starvation, or with small populations. Both large sample and very long study periods are necessary. For a year-long study in realistic conditions we may be trying to identify a change of the order of 400g in the average weight of the studied population and a change of 8 cal in their daily consumption. In other words, the very small population level changes in daily consumption and weight are truly emergent phenomena that cannot be observed at the individual level. Moreover, the huge variance in consumption and weight gain at the individual level casts doubt on the practicality of any “small changes” approach [44, 73] to interventions.

### Where Do All the Calories Go?

The above, observed weight differences are very small when compared to the expected weight gain if we posited the above population level energy imbalance, of the order of 200 cal, to be valid at the individual level, as noted in various more controlled, longitudinal studies [21–24] where, overfeeding specifically took place. For instance, in [35] a 75% increase in daily energy intake, corresponding to about 1,500 cal, for 20 days, led to an approximately 7% increase in weight (approx. 4kg) over that period, with a similar increase in Base Metabolic Rate (BMR). A naïve linear extrapolation would lead to a yearly weight increase of 80kg! For an excess consumption of 250 cals per day one would expect a yearly increase of 13.33kg. Indeed, taking the population level energy balance point to be at age 50, the excess calorie intake over the age period 20-50 is over one million calories! If one used the standard 3500 cal/pound rule this would correspond to an increase in weight of about 140kg over that time period. Even considering a much more sophisticated adaptive model [25–27] for relating excess consumption to weight gain, the actual, observed weight gain at the population level is far below what one would expect given the very large excess calorie intake indicated by the consumption pattern. In fact, just converting the overall weight gain at the population level into calories corresponds to a daily imbalance of about 7 cal; much, much smaller than our inferred energy imbalance of 200-300cal.

So, where do all these calories go? Our analysis dictates that this large excess translates into a relatively mild increase in BMI. Although it is generally accepted that obesity results from an energy imbalance between energy intake and energy expenditure, and metabolic energy expenditure can be measured relatively accurately using the doubly-labelled water technique, there is a great deal of uncertainty as to measurement of energy intake. Although many studies [20–24] have directly measured energy intake and expenditure in highly controlled, and hence highly artificial, settings, these are generally restricted to small populations and relatively short amounts of time. In these studies, energy intake can be measured in a relatively precise way, while in indirect studies energy intake is inferred via self-reported food intake, with the latter being subject to well-known reporting biases [53–58]. However, in both cases, there is a great deal of uncertainty as to how energy intake is partitioned among the different mechanisms of energy expenditure, such as base metabolic rate, thermogenesis, weight gain, movement, excretion etc.

To reconcile the apparent contradiction that a relatively large imbalance (approximately 200-300cal) in energy intake (consumption) seems to be driving a weight change that, energetically, could/should be explained by an imbalance of approximately 7 cal, we hypothesise that there are multiple physiological and behavioural mechanisms by which energy is dissipated and a higher degree of homeostasis than might be expected is maintained. Some of these are already known and have been discussed. The most obvious one is that an increase in weight leads to an increase in BMR. Using the Mifflin-St. Jeor equation the maximal year-on-year difference in BMR would be at age 20 and corresponds to a difference of only 7 cal. Indeed, over the 30-year period to BMI maximum the increase in BMR due to weight increase is largely cancelled by the reduction due to aging. Another factor would be physical activity, where it has been argued that heavier individuals will have higher energy requirements due to having to move a greater mass. However, in contrast to this is the gradual reduction in intense physical activity to be expected as a function of age.

Besides these potential factors, we believe there are potentially many more and that relative weight homeostasis is highly complex and adaptive and associated with multiple mechanisms. One, we believe to be particularly relevant is that of the effect of body, not core, temperature. In [75], in a pair of complementary experimental protocols, it was shown that body temperature increased linearly as a function of BMI, with an average difference of about 0.01 degrees Celsius per unit increment in BMI. Although this may seem very small it can correspond to a difference in energy loss of up to several hundred calories daily. Additionally, it has also been shown that the microbiota of an individual may play an important role [45, 46]. We believe that this role may potentially also be a positive one, reducing energy harvest as a potential adaptive response to an energy excess [47, 48].

Besides identifying potential adaptive mechanisms by which energy excess may be ameliorated, we believe it is also important to determine the characteristic timescale of each mechanism. For instance, it is clear that obesity itself at the population level is a long-term response to an energy imbalance. It may be that the relevant physiological and behavioural drivers of the obesity epidemic at such large scales are just not visible or easily measurable in short term experiments carried out under controlled conditions.

### Gender Differences and Unit Bias

Finally, we turn to the question of gender differences. Many studies have shown that women have a higher average BMI than men [44]. The data analysed here is consistent with this. However, we may go further and hypothesise that an important reason for this is that, in relative terms, the excess consumption of women is substantially higher than that for men. Of course, this begs the question of: why are women consuming in excess more than men? Besides a multitude of potential lifestyle factors, we wish to indicate a behavioural one associated with a particular cognitive bias – the “unit bias” [76]. This bias is such that people seem to think that a unit of some entity (with certain constraints) is the appropriate and optimal amount. Given that food portion sizes are generally gender neutral, it is clear that consumption of a portion will lead to an excess relative calorific intake that is greater for a woman than a man. If there is a bias towards consuming the entire portion for both men and women, then this would lead to a greater excess for women versus men.

## 5. Conclusions

There are several conclusions to be gleaned from our analysis that could and should have implications for public policy. Firstly, the strong convexity of the BMI curve as a function of age, which is seen in studies in other countries, indicates the importance of prevention measures in young adulthood, when considering the adult population. Various studies have shown the difficulty of weight reduction in the obese, for example [77], where only 5% of the obese managed to return to a normal weight. However, the challenge there is how to convince a young person who is currently only slightly overweight that they are on the path to obesity and therefore need to implement behaviour change now?

Secondly, the fact that the BMI curves are steeper for women than men, once again a feature common to several countries, indicates that young adult women are particularly susceptible to weight increase and therefore should be the target of specific interventions. We have argued that a potentially important contributing factor is the existence of the unit bias, in conjunction with the fact that portion sizes, especially outside the home, tend to be the same, independent of BMI, and therefore the consumption of the same portion by a woman or smaller man should have a relatively larger impact than on a large man.

Thirdly, the appearance of weight decrease beyond age 50, that accelerates with age, is consistent with many geriatric studies, where weight reduction can in fact be a health hazard. Although, weight reduction in the elderly is a multi-factorial phenomenon we argue that an important motor is simply an overall reduction in calorie consumption. Lastly, and perhaps more importantly, we have argued that an excess consumption that gradually decreases with age is an important motor for the obesity epidemic, with an inferred maximum excess of about 250 cal in young adults. Given that the energetically consistent imbalance associated with the observed weight gain is only about 7 cal, we argued that these two numbers can be reconciled by hypothesising the existence of multiple physiological and behavioural mechanisms by which weight homeostasis is maintained at an imbalance level consistent with a 7 cal per day imbalance in spite of the fact that the individual is assailed on average by an imbalance of up to 250 cal.

We suggested several possible mechanisms by which this enhanced homeostasis may be achieved, with associated evidence. An important potential implication of this is, if we invert the logic – inferring that, perhaps, a reduction of 250 cal per day over an extended period, up to 30 years, would only lead to a weight decrease equivalent to an 8 cal reduction! If this be the case, then this emphasizes the vital importance of preventing people from ever becoming overweight in the first place. In summary, the above conclusions can be stated as: You aren’t what you eat, you become what you eat - slowly.

### Study Limitations

The chief limitation of this study is that it is transverse not longitudinal. Although this does not affect the validity of our results it does play a role in their interpretation. In particular, in our positing a causal relationship between consumption and changes in BMI. A difference in BMI between two adjacent age groups, corresponding to two different populations, is interpreted as a BMI change within the same population. The chief possible source of error here is that adjacent age groups are statistically different with respect to potential confounding variables. As the study protocol was aimed at obtaining a representative sample over the Mexican population we believe that there are no significant differences in gender/educational level etc. between adjacent age groups. Of course, when comparing very distinct age groups, for example 20-21 with 79-80, there will be potentially significant differences. However, as our results are dependent on the consumption-BMI increase relation between adjacent age groups we believe that the significant differences that would accrue over a long time period will not affect our conclusions. For example, one might argue that the fact that the 80-year-old age group consume less than the 20-year-old age group is purely due to environmental factors, and therefore the interpretation that this represents an intrinsic reduction in an individual’s consumption as a function of age is incorrect. We believe it to be exceedingly unlikely that environmental factors have changed in precisely such a way so as to give rise to the behaviour seen in Figures 1-3 as opposed to these being principally a reflection of changes in an individual’s BMI and consumption as a function of age.

In the absence of detailed, long term, longitudinal studies that monitor consumption, inferences from transverse data are important from the point of view of hypothesis generation and self-consistency. It is also noteworthy that some of the base phenomenological elements, such as convexity of the BMI versus age relation and the decrease in calorie intake as a function of age, have also been noted in other large scales studies, such as NHANES.

Another limitation is that consumption is self-reported and, therefore, subject to the various biases examined at length in the literature [53–58]. Once again, in the absence of more precise measures applicable to large populations over long term periods, self-reporting remains the most important and used means to measure consumption.

A third limitation is that, in the absence of energy expenditure data, we are inferring an energy gap at the population level by relating self-reported consumption at the population level to weight gain at the population level.

## Data Availability

Data is available at: https://ensanut.insp.mx/encuestas/ensanut2006/index.php

## Acknowledgements

We are grateful for financial support from CONACyT Frontiers grant FC_2015- 2_1093 and DGAPA PAPIIT grant IG101520. We are also grateful for conversations with Katherine Stephens for suggesting the “unit bias” hypothesis.

## Supplementary material

Food items contained in each food category of the ENSANUT 2006 food frequency questionnaire.

| Vegetables: |  |  |  |
| --- | --- | --- | --- |
| Soup or broth with vegetables | Cream of vegetable soup | Tomato | Leafy greens |
| Chayote | Carrot | Zucchini | Broccoli or cauliflower |
| Cabbage | Green beans | Corn | Lettuce |
| Cactus leaves | Cucumber | Avocado | Other vegetables |
| Boiled potatoes | Nutritive drinks from the OPORTUNIDADES program |  |  |

| Milk products: |  |  |  |
| --- | --- | --- | --- |
| Liconsa milk | Other types of milk | Flavored milk | Cheese |
| Yoghurt | Danonino or similar | Yakult or similar |  |

| Cereals and grains: |  |  |
| --- | --- | --- |
| Peanuts, nuts and seeds | Pasta soup | Sopa de pasta b |
| Flour tortillas (MASECA or MINSA flour) | Tortillas (store bought) | Flour tortillas (wheat flour) |
| Lentils, garbanzos, broad bean, haricot bean | Beans a | Beans b |
| Tortillas made from nixtamal | Rice | White bread |
| Whole grain bread | Salted biscuits Galletas saladas | Breakfast cereals |

| Meat, fish, eggs: |  |  |  |
| --- | --- | --- | --- |
| Pork | Beef | Dried beef (machaca) | Sausage or chorizo |
| Chicken a | Chicken b | Chicken c | Egg a |
| Egg b | Fresh fish | Dried fish | Tuna and sardines |
| Seafood (shrimp, oysters, etc.) |  |  |  |
| Pork or turkey hotdog, ham or turkey slices etc. |  |  |  |

| Fast food: |  |  |  |
| --- | --- | --- | --- |
| Sandwiches | Hamburger | Pizza | Hot dog |
| Pozole | Tamal | Maize atole |  |
| Sopes, quesadillas, enchiladas, tacos or gorditas de comal (not fried) |  |  |  |
| Sopes, quesadillas, enchiladas, tacos or gorditas (fried) |  |  |  |

| Sugars and fats: |  |  |
| --- | --- | --- |
| Normal soft drink | Diet soft drink | Margerine |
| Juices and flavoured waters with sugar | Tea with sugar | Chocolate |
| Drinks with sugar | Coffee with sugar | Fried food |
| Milk with sugar | Cupcakes and doughnuts (indust.) | Cream |
| Milk with chocolate o other flavoring | Sweet biscuits | Butter |
| Sugar, for example in strawberries or bananas and cream | Jelly, Flan | Mayonnaise |
| Sweet bread (except doughnuts and churros) | Cereal bars | Sweets |
| Doughnuts and churro (bakery) | Cakes and pies | Fried banana |

| Fresh fruit: |  |  |  |
| --- | --- | --- | --- |
| Natural juices without sugar | Banana | Jicama | Orange or mandarin |
| Apple or pear | Watermelon or cantaloupe | Guayaba | Mango |
| Papaya | Pineapple | Grapefruit | Strawberry |
| Other fruits |  |  |  |

## References

1. World Health Organization. Obesity and Overweight Fact Sheet. World Health Organization; 2013. Available from: http://www.who.int/mediacentre/factsheets/fs311/en/index.html.

2. Mariel M Finucane, Gretchen A Stevens, Melanie J Cowan, Goodarz Danaei, John K Lin, Christopher J Paciorek, Gitanjali M Singh, Hialy R Gutierrez, Yuan Lu, Adil N Bahalim, Farshad Farzadfar, Leanne M Riley, Majid Ezzati, National, regional, and global trends in body-mass index since 1980: systematic analysis of health examination surveys and epidemiological studies with 960 country-years and 9.1 million participants, Lancet 2011; 377: 557–67.

3. Harry Rutter, The single most important intervention to tackle obesity…, Int J Public Health (2012) 57:657–658.

4. Michael Rosenbaum, Rudolph L. Leibel, and Jules Hirsch. Obesity, N Engl J Med 1997; 337:396–407

5. Jeffrey M. Friedman, Causes and control of excess body fat, Nature, Vol. 459 No. 21 May 2009

6. Sylvia R. Karasu, An Overview of the Complexities in Obesity: Limitations and Challenges, American Journal of Lifestyle Medicine, Vol 7, Issue 3, 2013

7. Sarah Frood & Lee M. Johnston & Carrie L. Matteson & Diane T. Finegood, Obesity, Complexity, and the Role of the Health System, Curr Obes Rep (2013) 2:320–326.

8. Anthony Fardet, Edmond Rock, From a Reductionist to a Holistic Approach in Preventive Nutrition to Define New and More Ethical Paradigms, Healthcare 2015, 3, 1054–1063.

9. Youfa Wang, Hong Xue, Layla Esposito, Michael J. Joyner, Yaneer Bar-Yam, and Terry T.-K. Huang, Applications of Complex Systems Science in Obesity and Noncommunicable Chronic Disease Research, Adv. Nutr. 5: 574–577, 2014; doi:10.3945/an.114.006650.

10. Ross A. Hammond, Chapter 61 - A Complex Systems Approach to Understanding and Combating the Obesity Epidemic, Editor(s): Laurette Dubé, Antoine Bechara, Alain Dagher, Adam Drewnowski, Jordan Lebel, Philip James, Rickey Y. Yada, Obesity Prevention, Academic Press, 2010, Pages 767–777.

11. David Albuquerque, Eric Stice, Raquel Rodríguez-López, Licíno Manco, Clévio Nóbrega, Current review of genetics of human obesity: from molecular mechanisms to an evolutionary perspective, Molecular Genetics and Genomics August 2015, Volume 290, Issue 4, pp 1191–1221.

12. Blanca M. Herrera, Sarah Keildson, Cecilia M. Lindgren, Genetics and epigenetics of obesity, Maturitas. 69(1): 41–49 (2011).

13. Boyd A Swinburn, Obesity prevention: the role of policies, laws and regulations, Australia and New Zealand Health Policy 2008, 5:12

14. Nicole L. Novak; Kelly D. Brownell, Role of Policy and Government in the Obesity Epidemic, Circulation. 2012;126: 2345–2352.

15. Ebbeling, C.B., Swain, J.F., Feldman, H.A. et al. 2012. “Effects of dietary composition on energy expenditure during weight-loss maintenance.” JAMA307(24): 2627–2634.

16. Kinsell, L.W., Gunning, B., Michaels, G.D., et al. 1964. “Calories do count.” Metabolism 13 (March): 195–204.

17. Andrea C Buchholz Dale A Schoeller, Is a calorie a calorie? The American Journal of Clinical Nutrition, Volume 79, Issue 5, 1 May 2004, Pages 899S–906S.

18. Nestle, M. and Nesheim, M. 2012. Why Calories Count: From Science to Politics. University of California Press.

19. Novotny, J.A., Gebauer, S.K., and D.J. Bauer. 2012. “Discrepancy between the Atwater factor predicted and empirically measured energy values of almonds in human diets.” American Journal of Clinical Nutrition 96: 296–301.

20. Keys A. The Biology of Human Starvation. University of Minnesota Press: Minneapolis, 1950.

21. John L. Sievenpiper, MD, PhD; Russell J. de Souza, ScD, RD; Arash Mirrahimi, HBSc; Matthew E. Yu, HBSc; Amanda J. Carleton, MSc; Joseph Beyene, PhD; Laura Chiavaroli, MSc; Marco Di Buono, PhD; Alexandra L. Jenkins, PhD, RD; Lawrence A. Leiter, MD; Thomas M.S. Wolever, MD, PhD; Cyril W.C. Kendall, PhD; David J.A. Jenkins, MD, PhD, DSc, Effect of Fructose on Body Weight in Controlled Feeding Trials: A Systematic Review and Meta-analysis, Ann Intern Med. 2012; 156(4):291–304.

22. Pao-Hwa Lin, Michael A Proschan, George A Bray, Claudia P Fernandez, Kimberly Hoben, Marlene Most-Windhauser, Njeri Karanja, Eva Obarzanek; Estimation of energy requirements in a controlled feeding trial, The American Journal of Clinical Nutrition, Volume 77, Issue 3, 1 March 2003, Pages 639–645, https://doi.org/10.1093/ajcn/77.3.639

23. Forbes, G. B., Brown, M. R., Welle, S. L., & Lipinski, B. A. (1986). Deliberate overfeeding in women and men: energy cost and composition of the weight gain. British Journal of Nutrition, 56(1), 1–9.

24. Webb P, Annis JF, Adaptation to overeating in lean and overweight men and women., Human Nutrition. Clinical Nutrition [01 Mar 1983, 37(2):117–131]

25. Hall, K. D., Sacks, G., Chandramohan, D., Chow, C. C., Wang, Y. C., Gortmaker, S. L., & Swinburn, B. A. (2011). Quantification of the effect of energy imbalance on bodyweight. The Lancet, 378(9793), 826–837.

26. Hall, K. D. (2010). Mechanisms of metabolic fuel selection: modeling human metabolism and body-weight change. IEEE Engineering in Medicine and Biology Magazine, 29(1), 36–41.

27. Hall KD. Computational model of in vivo human energy metabolism during semistarvation and refeeding. Am J Physiol Endocrinol Metab 2006; 291: E23–37.

28. National Health and Nutrition Examination Survey (NHANES), https://www.cdc.gov/nchs/nhanes/index.htm

29. Coronary Artery Risk Development in Young Adults (CARDIA) Study, https://www.cardia.dopm.uab.edu

30. Ashima K Kant and Barry I Graubard, Secular trends in patterns of self-reported food consumption of adult Americans: NHANES 1971–1975 to NHANES 1999–2002, Am J Clin Nutr. 2006; 84(5): 1215–1223.

31. Bray, G. A. (2007). The metabolic syndrome and obesity. Totowa, NJ: Humana Press.

32. Graham A Colditz, Walter C Willett, Meir J Stampfer, Stephanie J London, Mark R Segal, and Frank E Speizer, Patterns of weight change and their relation to diet in a cohort of healthy women, Am J Clin Nutr 1990; Sl: 1100-5.

33. Stefanie Vandevijvere, Carson C Chow, Kevin D Hall, Elaine Umali & Boyd A Swinburn, Increased food energy supply as a major driver of the obesity epidemic: a global analysis, Bull World Health Organ 2015;93:446–456.

34. Swinburn BA, Sacks G, Ravussin E. Increased food energy supply is more than sufficient to explain the US epidemic of obesity. Am J Clin Nutr. 2009; 90: 1453–1456.

35. Hall, K. D., Sacks, G., Chandramohan, D., Chow, C. C., Wang, Y. C., Gortmaker, S. L., & Swinburn, B. A. (2011). Quantification of the effect of energy imbalance on bodyweight. The Lancet, 378(9793), 826–837.

36. Schutz, Y., Byrne, N. M., Dulloo, A., & Hills, A. P. (2014). Energy gap in the aetiology of body weight gain and obesity: a challenging concept with a complex evaluation and pitfalls. Obesity facts, 7(1), 15–25.

37. Wang, Y. C., Gortmaker, S. L., Sobol, A. M., & Kuntz, K. M. (2006). Estimating the energy gap among US children: a counterfactual approach. Pediatrics, 118(6), e1721–e1733.

38. Bouchard, C. (2008) The magnitude of the energy imbalance in obesity is generally underestimated, International journal of obesity, 32(6), 879.

39. Butte, N. F., Christiansen, E., & Sørensen, T. I. (2007). Energy imbalance underlying the development of childhood obesity. Obesity, 15(12), 3056–3066.

40. Bray, G. A., & Champagne, C. M. (2005). Beyond energy balance: there is more to obesity than kilocalories. Journal of the American Dietetic Association, 105(5), 17–23.

41. Millward DJ. Energy balance and obesity: a UK perspective on the gluttony v. sloth debate. Nutr Res Rev. 2013; 26:1–21.

42. Hill JO. Understanding and addressing the epidemic of obesity: an energy balance perspective. Endocr Rev. 2006 Dec; 27:750–761.

43. Hill JO, Peters JC, Wyatt HR. Using the energy gap to address obesity: a commentary. J Am Diet Assoc. 2009;109:1848–1853.

44. Hill JO, Wyatt HR, Reed GW, Peters JC. Obesity and the environment: where do we go from here? Science. 2003; 299:853–5.

45. Turnbaugh, P. J. and Gordon, J. I. (2009), The core gut microbiome, energy balance and obesity. The Journal of Physiology, 587: 4153–4158. doi:10.1113/jphysiol.2009.174136

46. Reiner Jumpertz, Duc Son Le, Peter J Turnbaugh, Cathy Trinidad, Clifton Bogardus, Jeffrey I Gordon, and Jonathan Krakoff, Energy-balance studies reveal associations between gut microbes, caloric load, and nutrient absorption in humans, Am J Clin Nutr 2011; 94:58–65.

47. Lin, H. V., Frassetto, A., Kowalik Jr, E. J., Nawrocki, A. R., Lu, M. M., Kosinski, J. R., … & Marsh, D. J. (2012). Butyrate and propionate protect against diet-induced obesity and regulate gut hormones via free fatty acid receptor 3-independent mechanisms. PloS one, 7(4), e35240.

48. Conterno, L., Fava, F., Viola, R., & Tuohy, K. M. (2011). Obesity and the gut microbiota: does up-regulating colonic fermentation protect against obesity and metabolic disease? Genes & nutrition, 6(3), 241.

49. A. E. Black, W.A. Coward, T.J. Cole and A.M. Prentice, Human energy expenditure in affluent societies: an analysis of 574 doubly-labelled water measurements, European Journal of Clinical Nutrition 1996, 50(2):72–92.

50. R.J. Hill and P.S.W. Davies, The Validity of self-reported energy intake as determined using the doubly labelled water technique, Brit. J. Nutr. 2001, 85 (4): 415–430.

51. National Institute for Public Health (Mexico). Mexico National Survey of Health and Nutrition 2005-2006. Cuernavaca, Mexico: National Institute for Public Health (Mexico)

52. Olaiz-Fernández G, Rivera-Dommarco J, Shamah-Levy T, Rojas R, Villalpando-Hernández S, Hernández-Avila M, Sepúlveda-Amor J. Encuesta Nacional de Salud y Nutrición 2006. First Edition Cuernavaca, México: Instituto Nacional de Salud Pública, 2006.

53. Macdiarmid, J., & Blundell, J. (1998) Assessing dietary intake: Who, what and why of under-reporting, Nutrition Research Reviews, 11 (2), 231–253.

54. Johnson, R. K. (2002), Dietary Intake—How Do We Measure What People Are Really Eating? Obesity Research, 10: 63S-68S. doi:10.1038/oby.2002.192

55. H. Schröder, M.I. Covas, J. Marrugat, J. Vila, A. Pena, M. Alcántara, R. Masiá, Use of a three-day estimated food record, a 72-hour recall and a food-frequency questionnaire for dietary assessment in a Mediterranean Spanish population, Clinical Nutrition, Volume 20, Issue 5, 2001, Pages 429-437, ISSN 0261-5614.

56. Anja Kroke, Kerstin Klipstein-Grobusch, Susanne Voss, Jutta Möseneder, Frank Thielecke, Rudolf Noack, Heiner Boeing; Validation of a self-administered food-frequency questionnaire administered in the European Prospective Investigation into Cancer and Nutrition (EPIC) Study: comparison of energy, protein, and macronutrient intakes estimated with the doubly labeled water, urinary nitrogen, and repeated 24-h dietary recall methods, The American Journal of Clinical Nutrition, Volume 70, Issue 4, 1 October 1999, Pages 439–447.

57. Fernanda B. Scagliusi, Eduardo Ferriolli, Karina Pfrimer, Cibele Laureano, Caroline Sanita Cunha, Bruno Gualano, Barbara Hatzlhoffer Lourenço, Antonio Herbert Lancha, Underreporting of Energy Intake in Brazilian Women Varies According to Dietary Assessment: A Cross-Sectional Study Using Doubly Labeled Water, Journal of the American Dietetic Association, Volume 108, Issue 12, 2008, Pages 2031–2040.

58. Cade, J., Thompson, R., Burley, V., & Warm, D. (2002) Development, validation and utilisation of food-frequency questionnaires – a review, Public Health Nutrition, 5(4), 567–587.

59. Pérez Lizaur, A. B., Palacios González, B., Castro Becerra, A. L., & Flores Galicia, I. (2008). Sistema mexicano de alimentos equivalentes. México.

60. Mifflin, M. D., St Jeor, S. T., Hill, L. A., Scott, B. J., Daugherty, S. A., & Koh, Y. O. (1990). A new predictive equation for resting energy expenditure in healthy individuals. The American journal of clinical nutrition, 51(2), 241–247.

61. United States Department of Agriculture (USDA), 2014. “National Nutrient Database for Standard Reference” USDA Viewed: 29th April 2015. http://ndb.nal.usda.gov/ndb/search/list

62. Svensson, E., Reas, D. L., Sandanger, I., & Nygård, J. F. (2007). Urban—rural differences in BMI, overweight and obesity in Norway (1990 and 2001). Scandinavian journal of public health, 35(5), 555–558.

63. Reas, D. L., Nygård, J. F., Svensson, E., Sørensen, T., & Sandanger, I. (2007). Changes in body mass index by age, gender, and socio-economic status among a cohort of Norwegian men and women (1990–2001). BMC public health, 7(1), 269.

64. Judith J. Wurtman Harris Lieberman Rita Tsay Tony Nader Beverly Chew, Calorie and Nutrient Intakes of Elderly and Young Subjects Measured Under Identical Conditions Journal of Gerontology, Volume 43, Issue 6, 1 November 1988, Pages B174–B180, https://doi.org/10.1093/geronj/43.6.B174

65. Robert R. Wolfe, Sharon L. Miller, Kevin B. Miller, Optimal protein intake in the elderly, Clinical Nutrition, Volume 27, Issue 5, 2008, Pages 675–684,

66. Bruno Vellas, Yves Guigoz, Philip J Garry, Fati Nourhashemi, David Bennahum, Sylvie Lauque, Jean-Louis Albarede, The mini nutritional assessment (MNA) and its use in grading the nutritional state of elderly patients, Nutrition, Volume 15, Issue 2, 1999, Pages 116–122.

67. C Castaneda, J M Charnley, W J Evans, M C Crim; Elderly women accommodate to a low-protein diet with losses of body cell mass, muscle function, and immune response, *The American Journal of Clinical Nutrition*, Volume 62, Issue 1, 1 July 1995, Pages 30–39

68. Ahmed, Tanvir, and Nadim Haboubi. “Assessment and Management of Nutrition in Older People and Its Importance to Health.” Clinical Interventions in Aging 5 (2010): 207–216.

69. Sheiham, A., & Steele, J. (2001) Does the condition of the mouth and teeth affect the ability to eat certain foods, nutrient and dietary intake and nutritional status amongst older people? Public Health Nutrition, 4(3), 797–803.

70. Brownie, S. (2006), Why are elderly individuals at risk of nutritional deficiency? International Journal of Nursing Practice, 12: 110–118.

71. John R. Speakman et al, (2011) “Set points, settling points and some alternative models: theoretical options to understand how genes and environments combine to regulate body adiposity, “Dis Model Mech. 4(6): 733–745.

72. Swinburn, B. A., Sacks, G., Lo, S. K., Westerterp, K. R., Rush, E. C., Rosenbaum, M., … & Ravussin, E. (2009). Estimating the changes in energy flux that characterize the rise in obesity prevalence–. The American journal of clinical nutrition, 89(6), 1723–1728.

73. Hill, J. O. (2008). Can a small-changes approach help address the obesity epidemic? A report of the Joint Task Force of the American Society for Nutrition, Institute of Food Technologists, and International Food Information Council. The American journal of clinical nutrition, 89(2), 477–484.

74. Ulf Ekelund, Jan Åman, Agneta Yngve, Cecilia Renman, Klaas Westerterp, Michael Sjöström; Physical activity but not energy expenditure is reduced in obese adolescents: a case-control study, The American Journal of Clinical Nutrition, Volume 76, Issue 5, 1 November 2002, Pages 935–941.

75. Fossion, R., Stephens, C. R., García-Pelagio, K. P., & García-Iglesias, L. (2017, July). Data Mining and Time-Series Analysis as Two Complementary Approaches to Study Body Temperature in Obesity. In Proceedings of the 2017 International Conference on Digital Health (pp. 190–194). ACM.

76. Geier, A. B., Rozin, P., & Doros, G. (2006). Unit bias: A new heuristic that helps explain the effect of portion size on food intake. Psychological Science, 17(6), 521–525.

77. Fildes, A., Charlton, J., Rudisill, C., Littlejohns, P., Prevost, A. T., & Gulliford, M. C. (2015). Probability of an obese person attaining normal body weight: cohort study using electronic health records. American Journal of Public Health, 105(9), e54–e59.

